# Anterior cingulate sulcation is associated with onset and survival in frontotemporal dementia

**DOI:** 10.1101/2023.03.30.23287945

**Authors:** Luke Harper, Sterre de Boer, Olof Lindberg, Jimmy Lätt, Nicholas Cullen, Lyles Clark, David Irwin, Lauren Massimo, Murray Grossman, Oskar Hansson, Yolande Pijnenburg, Corey T. McMillan, Alexander F Santillo

## Abstract

**Background:** Frontotemporal dementia is the second most common form of early onset dementia (< 65 years). Despite this there are few known disease modifying factors. The anterior cingulate is a focal point of pathology in behavioural variant frontotemporal dementia. Sulcation of the anterior cingulate is denoted by the presence of a paracingulate sulcus, a tertiary sulcus developing, where present during the third gestational trimester and remaining stable throughout life. This study aims to examine the impact of right paracingulate sulcal presence on the expression and prognosis of behavioural variant Frontotemporal Dementia.

**Methods:** This retrospective analysis drew it’s population from two clinical samples recruited from memory clinics at University Hospitals in The United States of America and The Netherlands. Individuals with sporadic behavioural variant Frontotemporal Dementia were enrolled between 2004 and 2022 and followed up for an average of 7.71 years. T1-MRI data were evaluated for hemispheric paracingulate sulcal presence in accordance with an established protocol by two blinded raters. Outcome measures included age at onset, survival, cortical thickness, and Frontotemporal Lobar Degeneration-modified Clinical Dementia Rating determined clinical disease progression.

**Results:** The study population consisted of 186 individuals with sporadic behavioural variant Frontotemporal Dementia, (113 males and 73 females) mean age 63.28 years (SD 8.32). The mean age at onset was 2.44 years later in individuals possessing a right paracingulate sulcus (60.2 years (SD 8.54)) versus individuals who did not (57.76 (8.05)), 95% CI >0.41, *P* = 0.02. Education was not associated with age at onset (β = -0.05, *P*=0.75). Presence of a right paracingulate sulcus was associated with a 119% increased risk of death per year after age at onset (HR 2.19, CI [1.21 - 3.96], *P*<0.01), whilst the mean age at death was similar for individuals with a present and absent right paracingulate sulcus (*P* = 0.7). Right paracingulate sulcal presence was not associated with baseline cortical thickness.

**Conclusion:** Right paracingulate sulcal presence is associated with disease expression and survival in sporadic behavioural variant Frontotemporal Dementia. Findings provide evidence of neurodevelopmental brain reserve in behavioural variant Frontotemporal Dementia which may be important in the design of trials for future therapeutic approaches.

## Introduction

Behavioural variant Frontotemporal Dementia (bvFTD) is the most common clinicoradiological syndrome within the Frontotemporal Dementias (FTD). FTD is highly heritable, approximately 30% of suffers have a strong family history with heritability accounted for in 10-20% of FTD by autosomal dominant mutations in the *chromosome 9 open reading frame 7*2 (*C9orf72*), *progranulin* (*GRN*), or *microtubule-associated protein tau* (*MAPT*) genes.^1, 2^ The mean age at onset (AAO) is between 45 and 65 years.^3^ There are however documented cases under the age of 30, whilst up to 30% of patients have a later onset (≥65 years).^3^ AAO is variably affected by pathological genetic mutations, where present and genetic variation, including the presence of risk allele rs1990622 in the *TMEM106B* gene.^4-6^ Despite these few exceptions there are no known factors affecting AAO in bvFTD. Environmental disease modifying factors, including educational, occupational attainment and occupational engagement have been shown to provide resilience to the neuropathological burden of FTD, however they have yet to be shown to be associated with AAO.^7 8 5, 9-11^ BvFTD atrophy has a predilection for the Anterior Cingulate (AC) and frontoinsula regions.^12^ Gyrification of the AC may be characterised morphologically by the presence of a paracingulate sulcus (PCS), a tertiary sulcus which, where present develops during the third trimester of gestation denoting the existence of a Paracingulate Gyrus (PCG).^13^ Paracingulate sulcation is more frequent in the left hemisphere of healthy individuals.^14-17^ The PCG is active during performance of cognitively demanding tasks drawing on higher-order executive function where possession of leftward PCS asymmetry (presence of a left but not right hemisphere PCS, as displayed in **Fig. 1)** has been associated with a performance advantage.^18 19^ Furthermore individuals with asymmetric PCS patterns display greater inhibitory control and cognitive efficiency.^20 21 22 23^ Conversely, in schizophrenia, a reduced distribution of leftward PCS asymmetry is observed and interpreted as evidence of a prenatal neurodevelopmental aberration in the pathogenesis of Schizophrenia.^15, 17^

**Fig. 1.**
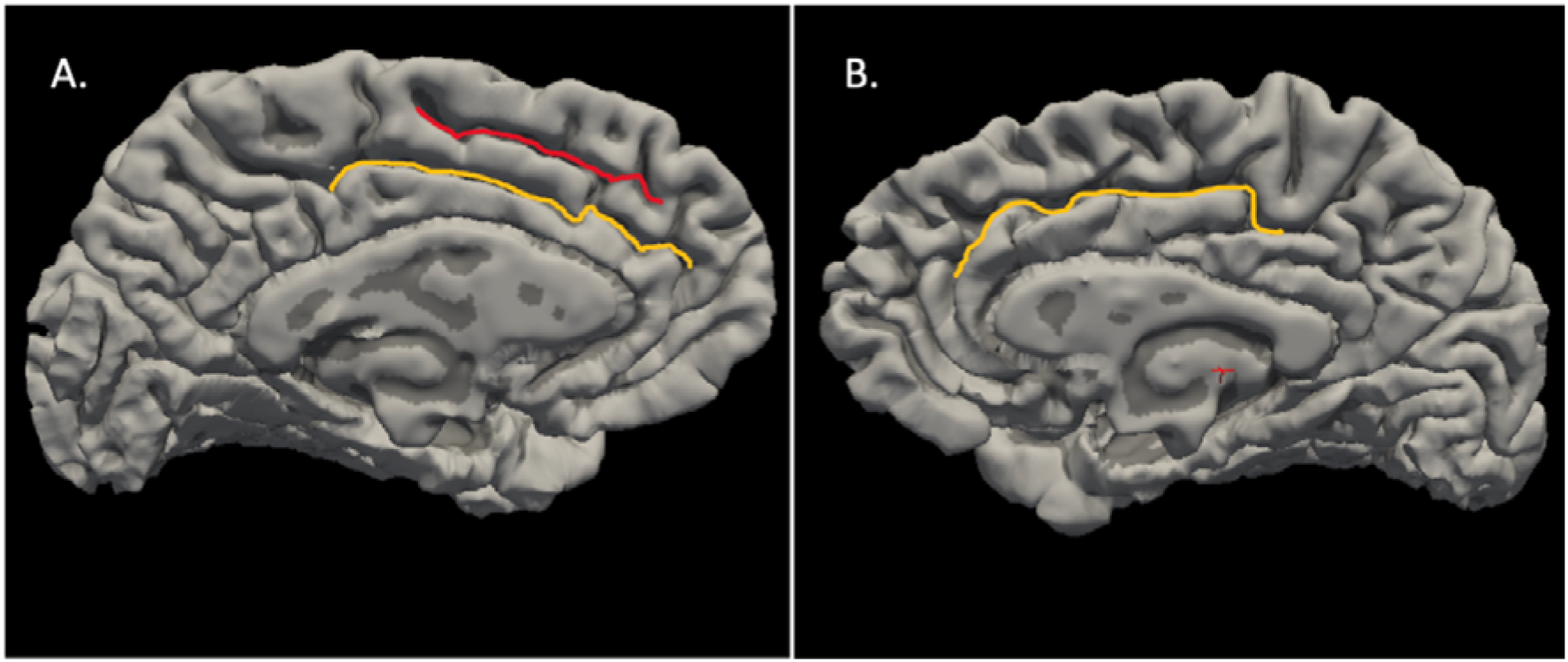
Cingulate and Paracingulate Sulci Identification and Measurement. 58-year-old male with probable bvFTD displays a Leftward pattern of paracingulate asymmetry. A. Sagittal image of the pial surface of the left hemisphere displaying a prominent, (length ≥40mm) left paracingulate sulcus (red) and the cingulate sulcus (yellow). B. Sagittal image of the pial surface of the right hemisphere displaying the cingulate sulcus (yellow) with absence of a right PCS.

In previous work the distribution of hemispheric PCS frequency in sporadic bvFTD was similar to that of healthy individuals.^24^ Significantly however, right PCS presence was associated with a later AAO in sporadic bvFTD, and is a potential proxy of brain reserve.^24^ Studied proxies of reserve provide resilience to disease burden prior to phenoconversion, following this critical point compensatory reserve mechanisms become overwhelmed and clinical decline proceeds more rapidly than in those lacking such proxies.^25^ Despite the possible effect on disease expression the impact of right PCS presence on disease progression and survival after AAO in bvFTD is not yet known. The present study aims to confirm and expand upon findings from Harper et al 2022^24^ in a novel independent cohort with longitudinal data. Our primary hypothesis was an association between right PCS presence and a later AAO in sporadic bvFTD. Secondary hypotheses were that following disease onset, and independent of education, disease progression would occur more rapidly, and survival would be shorter in individuals possessing a right PCS. Our tertiary hypothesis was that whilst at the same clinical stage, bvFTD individuals with a present right PCS would display greater disease burden, demonstrated by cortical atrophy, than those without.

## Materials and Methods

### Participants

This retrospective analysis included individuals with sporadic bvFTD drawn from two clinical samples recruited from memory clinics at university hospitals in the United States of America (Penn FTDC, enrolment between 2004 and 2018) and The Netherlands (Amsterdam Dementia Cohort,^26^ enrolment between 2000 to 2022). BvFTD was diagnosed in accordance with revised International bvFTD Consortium criteria^27^ following multidisciplinary team assessment, clinical examination, standardized symptom assessment, neuropsychological and neurological examination, blood, and cerebrospinal fluid analysis of core Alzheimer’s Disease (AD) biomarkers and brain MRI. Detailed cohort descriptions are published in the **Supplement**. Individuals with suspected hereditary bvFTD; Woods^28^ criteria “High” or “Medium” (Penn FTDC) or Goldmans^29^ criterion ≤ 3.0 (Amsterdam Dementia Cohort) were excluded. *C9orf72* mutations were excluded in all and *GRN* and *MAPT* mutations were excluded in 92 individuals. Two individuals met subclassification criteria^30^ for right temporal lobe variant FTD and eight individuals met FTD-ALS criteria^31^ and were excluded. Neuropathological data was available in 38 individuals. Where neuropathology was consistent with isolated non-FTLD neurodegenerative disease, (n=4) individuals were excluded whereas individuals with concomitant FTLD and non-FTLD neurodegenerative disease were included. All subjects gave informed consent in accordance with the Declaration of Helsinki prior to inclusion in their native studies. Native studies were conducted with approval of respective local ethics committees, as detailed in the **Supplement**.

### Magnetic Resonance Image Acquisition and Software

High-resolution volumetric whole brain T1-weighted magnetic resonance (MR) images were obtained from all individuals using 1.5 or 3.0 Tesla systems with a minimum spatial resolution of 1.5□×□1.5□×□1.5 mm. Protocols and MRI related details are provided in the **Supplement**.

Prior to analysis, images were pseudo-anonymized and visually inspected. Six individuals were removed due to distortion of their MR data by movement artifact. A further two individuals were removed with postoperative intracranial anatomy obscuring PCS identification. Cortical reconstruction and volumetric segmentation were performed on T1 3D MR images using FreeSurfer Software version 7.3.2 image analysis pipeline, (http://surfer.nmr.mgh.harvard.edu/). This procedure is described elsewhere^32^ and briefly in the **Supplement**. Reconstructed data sets were visually inspected for accuracy by a single rater. Cortical thickness was successfully calculated in 176 individuals. Nineteen studies were excluded following quality control on the basis of poor surface reconstruction.

### Paracingulate Sulcus Measurement and Classification Criteria

Manual PCS classification was performed radiographically according to a protocol adapted from Garrison’s established protocol for PCS classification^33^, which has been used and described in Harper et al 2022^24^ and is documented in full in the **Supplement**. The PCS was categorized in a binary fashion; “present” (≥20 mm) or “absent” (<20 mm), as is standard amongst PCS classification protocols.^15, 17, 33-35^ Additionally, present PCS were subclassified as “prominent” where their length exceeded 40mm, as performed by others.^33 14^ Sulcation ratings were performed by two raters, LH and AS, who were blinded to individuals’ clinical and demographic data.

### Clinical Disease Expression, Progression and Survival

AAO was determined by a clinician based on patient and caregiver history as the first date at which typical symptoms, compatible with a diagnosis of bvFTD^27^ became apparent. Disease severity was assessed longitudinally in the Penn FTDC cohort at baseline and follow-up according to the Frontotemporal Lobar Degeneration-modified Clinical Dementia Rating (FTLD-CDR)^36^ The FTLD-CDR was selected over other clinical rating tools due to its superiority in the accurate classification of disease severity in FTD.^36, 37^ Survival data was recorded in all individuals.

### Statistical Analysis

Group differences in categorical variables were tested using Chi-Squared tests. Continuous measures were compared using two-sample t-tests with normality tested by the Shapiro-Wilk test. A one-sided *t*-test was conducted to analyse the association between right PCS presence and AAO. Effect sizes were calculated according to Cohen’s d. Simple and multiple linear regression models were fitted to evaluate covariable and interaction effects on AAO. Correlations between continuous variables were calculated using Pearson’s method whilst correlation between categorical and continuous variables utilised point biserial correlation. Survival analyses were performed from AAO to time of death from any cause (outcome = 1) or censoring date (outcome = 0). The censoring date was recorded as the date of last contact with the individual. Survival analyses were carried out using the Kaplan-Meier method with log rank post hoc testing by means of univariate and multivariate stepwise Cox proportional-hazard regression analysis. Hazard Ratios (HR) are provided with 95% confidence intervals (CIs). A linear mixed effects model with random intercepts and slopes was fitted to analyse clinical disease progression. Cortical thickness calculations were undertaken in FreeSurfer Software version 7.3.2 (http://surfer.nmr.mgh.harvard.edu/). Group differences in cortical thickness according to right PCS presence were analysed by fitting a vertex-based general linear model corrected for the effect of age and sex. Cluster-wise correction for multiple comparisons was performed using Monte Carlo simulation,^38^ with a threshold of *P*<0.05. Statistical analysis was performed using R software (R CoreTeam 2016, https://www.r-project.org/). *P*<0.05 was considered statistically significant. Statistical procedures related to primary and secondary hypotheses alongside power calculations were pre-registered and may be accessed at https://aspredicted.org/SKM_6C1.

## Data Availability

Anonymized data will be shared by request from a qualified academic investigator for the sole purpose of replicating procedures and results presented in the article if data transfer is in agreement with relevant legislation on the general data protection regulation and decisions and by the relevant Ethical Review Boards, which should be regulated in a material transfer agreement.

## Results

The study population consisted of 186 sporadic bvFTD individuals, (113 males and 73 females) with a mean age of 63.28 years (SD 8.32) and AAO of 59.15 (8.4). Demographic and results data are displayed in **Table 1** and the **Supplement**.

**Table 1.** Study Population & Paracingulate Status.

|  | Entire Population | Penn FTDC | Amsterdam Dementia Cohort | P-value |
| --- | --- | --- | --- | --- |
| Participants | 186 | 94 | 92 |  |
| Age, mean (SD), years | 63.28 (8.32) | 63.15 (8.82) | 63.38 (7.98) | 0.87 |
| Age at Onset, mean, (SD), years | 59.15 (8.4) | 58.16 (8.29) | 60.14 (8.43) | 0.11 |
| Sex | 113M: F73 | 59M: 35F | 54M: 38F | 0.54 |
| Education,<sup>a</sup> years (SD) | 13.34 (3.79) | 15.88 (2.74) | 10.63 (2.72) | <000.1 |
| Handedness, No. |  |  |  | 0.69 |
| Right | 146 | 81 | 65 |  |
| Left | 12 | 9 | 3 |  |
| Ambidextrous | 5 | 4 | 1 |  |
| Unknown | 23 | 0 | 23 |  |
| Diagnostic classification,<sup>b</sup> No |  |  |  |  |
| Possible | 8 | 8 | 0 |  |
| Probable | 140 | 51 | 89 |  |
| Definite | 26 | 24 | 2 |  |
| Unknown | 11 | 11 | 0 |  |
| MMSE, mean (SD) | 24.18 (5.35) | 23.63 (6.07) | 24.8 (4.35) | 0.14 |
| FTLD CDR Sum of boxes, mean (SD) | 7.51 (3.83) | 9.01 (3.74) | 6.71 (3.65) | 0.001 |
| FTLD CDR Global scores, mean (SD) | 1.59 (0.72) | 1.79 (0.59) | 1.45 (0.76) | 0.01 |
| Hemispheric PCS, No. (%) |  |  |  |  |
| Left present | 145 (78) | 73 (78) | 72 (78) | 1 |
| Left prominent | 59 (32) | 28 (30) | 31 (34) | 0.68 |
| Right present | 106 (57) | 58 (62) | 48 (52) | 0.53 |
| Right prominent | 31 (17) | 18 (19) | 13 (14) | 0.47 |
| Deceased,<sup>c</sup> No. (%) | 112 (60) | 51 (56) | 61 (66) | 0.13 |
| Age at Death, mean (SD), years | 67.19 (8.86) | 67.42 (9.14) | 66.92 (8.62) | 0.79 |

Demographic and Results data for entire population and according to cohort, Penn FTDC (The University of Pennsylvania Frontotemporal Degeneration Centre, Pennsylvania, USA) and Alzheimer center of the VU University Medical Center, Amsterdam (Amsterdam Dementia Cohort).<sup>26</sup>
Standard deviation (SD). Sex data: Male (M). Female (F). <sup>a</sup> Education, years data available for 94/94 Penn FTDC and 88/92 Amsterdam Dementia Cohort. <sup>b</sup> Diagnostic classification according to revised International bvFTD Consortium criteria.<sup>27</sup> <sup>c</sup> Survival data was available for 185/186 individuals. Mini Mental State Examination (MMSE). Frontotemporal Lobar Degeneration-modified Clinical Dementia Rating (FTLD-CDR)<sup>36</sup>, Sum of boxes (Sum) and Global score (Global). Hemispheric Paracingulate Sulcus (Hemispheric PCS), present = PCS length $\geq 20$ mm, prominent = PCS length $\geq 40$ mm. Frequency. T-tests and Chi-Squared tests were performed to evaluate differences in continuous and nominal data, respectively.

Mean AAO was 2.44 years later (Cohen’s d = 0.29, CI [>0.41], *P*=0.02) in individuals with a present (mean AAO 60.20 years (SD 8.54)) versus absent right PCS (57.76 (8.05)). These data are displayed in **Fig. 2**. Independently both the Penn FTDC (Mean Difference (MD) = 2.48 years, *P*=0.08) and the Amsterdam Dementia Cohort (MD = 2.8 years, *P*=0.06) showed a trend towards greater AAO in individuals with a present right PCS. Importantly this analysis was powered to identify significance at n = 173. Mean AAO did not differ significantly according to left PCS presence, (MD = 1.53, *P*=0.16). Right PCS length was not significantly correlated with AAO (r = 0.04, CI [-0.15 – 0.23], *P*=0.66) however mean AAO in individuals with a prominent right PCS was 3.11 years later than in individuals with an absent right PCS, (Mean AAO = 60.87 years (SD 6.92) and 57.76 (8.05), respectively, (t = 2.02, CI [>0.55], *P*=0.02). Education was similar in individuals with a present and absent right PCS (t = 1.12, CI [-0.5-1.82], *P*=0.26). A correlation between education and AAO was not observed, (*r*□=□0.09, CI [-0.06–0.23], *P*=0.25). In linear models neither education, sex or handedness were independently associated with AAO, (*P*=0.75, 0.3 and 0.45 respectively) nor was an association with AAO identified after adjusting for right PCS presence, (*P*=0.64, 0.41 and 0.27 respectively). Furthermore, inclusion of these variables failed to improve the overall predictability of these models. An interaction effect between education (β = 0.05, CI [-0.6 – 0.71], *P*=0.87) and right PCS presence on AAO was not identified. Sex however interacted with right PCS presence such that AAO was greater in males processing a right PCS, (β = 5.14, CI [0.17 – 10.1], *P*=0.04), **Supplementary Fig. 1**.

**Fig. 2.**
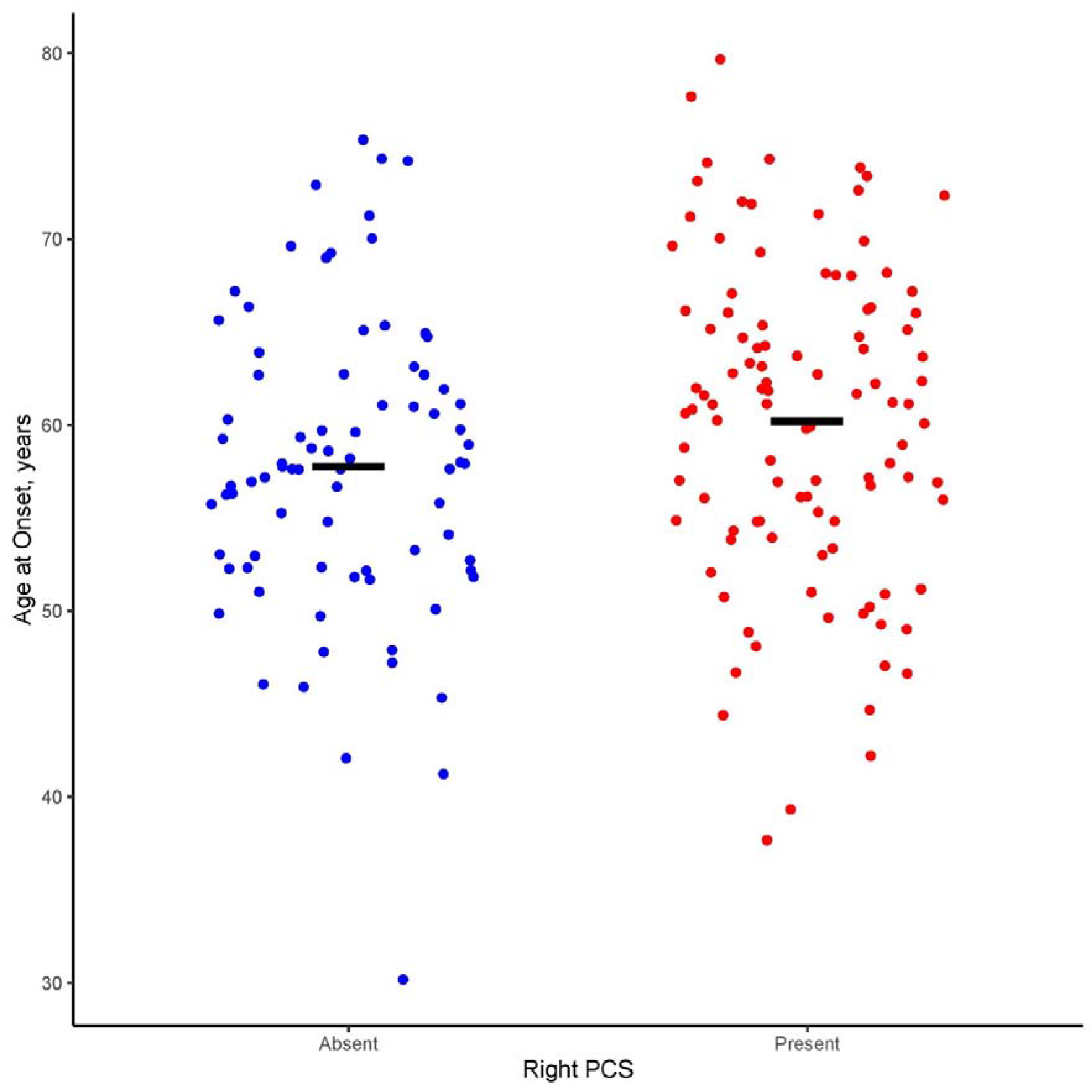
Age at symptom onset by Right Hemisphere Paracingulate Sulcal presence in bvFTD. Red dots represent individuals with a present right Paracingulate Sulcus (PCS), n=80. Blue dots represent individuals with an absent right PCS, n=106. Black lines represent group means of age at symptom onset; 57.76 (SD 8.05) and 60.2 (8.54) respectively for individuals with an absent and present right PCS, mean difference = 2.44, *P*=0.02.

Individuals were followed for a median of 7.71 years (IQR 5.00 - 10.8.7). Mean age at death was similar in individuals with present (66.94 years, (SD 9.66) and absent (67.62, (7.38)) right PCS, *P*=0.7. Survival was significantly affected by right PCS presence, (Chi-squared 6.6, *P*=0.01). The unadjusted risk of death per year after AAO was 65.1% greater in individuals processing a right PCS (HR 1.65, CI [1.13-2.42], *P*=0.01). Kaplan Meier estimates for this result are presented in **Fig. 3**. Risk of death was enhanced to 119% following correction for baseline FTLD-CDR, AAO, handedness, sex, and years of education (HR 2.19, CI [1.21-3.96], *P*<0.01). Sensitivity analyses after fitting this model identified a significantly increased risk of death following AAO in both the Amsterdam Dementia Cohort (HR 2.38, CI [1.09–5.16], *P*=0.029) and the Penn FTDC cohort (HR 3.83, CI [1.05–14.03], *P*=0.043). No interaction effect of sex and right PCS presence on survival was observed, (*P*>0.1).

**Fig. 3.**
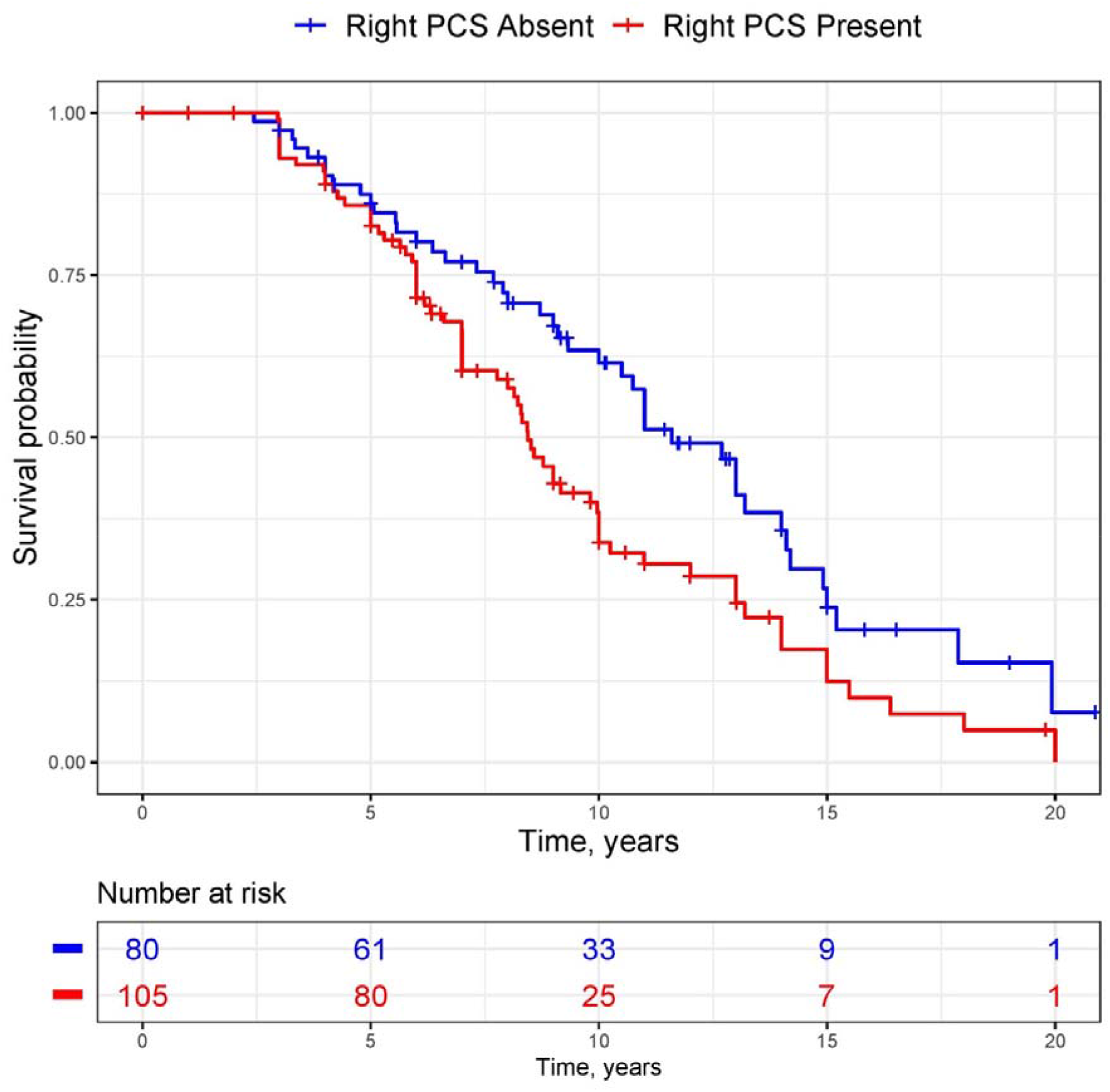
Kaplan-Meier Estimate of Survival by Right Paracingulate Sulcal presence. Kaplan Meier curve for survival in individuals with behavioral variant Frontotemporal Dementia (n=185) according to right paracingulate sulcal presence. Red line indicates survival in individuals with a present right Paracingulate Sulcus (PCS). Blue line represents individuals with an absent right PCS.

Longitudinal FTLD-CDR data were available for 49 individuals with a median follow-up time of 15.83 months. FTLD-CDR sum of boxes and global scores at baseline were similar in individuals with present and absent right PCS after adjusting for age, handedness, sex, and education, (β=-1.23, CI [-0.59–0.14], *P*=0.08 and β=-0.22, CI [-0.48–0.04], *P*=0.1 respectively). In a linear mixed effects model with covariates; age, sex, education, and handedness there was suggestion of a faster rate of clinic disease progression in individuals possessing a right PCS though results did not reach statistical significance, (FTLD-CDR sum of boxes; β=1.01, CI [-0.65-2.63], *P*=0.23 and FTLD-CDR global score; β=0.09, CI [-0-.11-0.29], *P*=0.36). These data are depicted in **Supplement Fig. 2**.

Individuals were at a similar clinical disease stage at time of MRI imaging, with no association identified between baseline FTLD-CDR Sum of boxes scores and right PCS presence, corrected for time from MRI imaging to CDR-FTLD scoring (β=-0.71, CI [-1.99-0.57], *P*=0.27). Twenty-three regions of interest were initially identified with significant differences in cortical thickness according to right PCS presence. All however failed to survive cluster correction for multiple analysis. Cluster details are reported in the **Supplement**, Table 2 and 3.

## Discussion

In keeping with Harper et al 2022,^24^ this study provides confirmation in a novel, adequately powered cohort of an association between right PCS presence and a later AAO in sporadic bvFTD. Moreover, we demonstrate that this effect is independent of education and that right PCS presence is a prognostic biomarker in sporadic bvFTD, associated with worse survival following AAO. These findings have important consequences; they develop our understanding of the natural history of sporadic bvFTD, give an insight into the implications of neurodevelopmental variability on the expression of a neurodegenerative disease and offer a proxy for brain reserve in bvFTD with potential implications for therapeutic trials.

Reserve theories suggest that individuals possessing adaptable functional brain processes; cognitive reserve, and/or preferential neurobiological capital; brain reserve, possess resilience to the clinical manifestations of a disease despite significant pathological burden.^39^ Cognitive reserve is a widely accepted concept in AD^40 41 42, 43^ where lifetime experiences, including and most well described but not limited to educational attainment is associated with reduced age-specific risk of developing AD.^39, 44, 45^ Furthermore, the concept of motor reserve has recently emerged in Parkinson’s Disease.^46^ In FTD greater occupational attainment,^7^ degree of occupational^8^ and active leisure engagement^47^ have, in some studies, been associated with increased functional and/or structural cerebral impairment despite comparable clinical severity. Furthermore, individuals with greater composite educational and occupational attainment scores process increased brain maintenance, with preservation of frontal anatomical integrity compared to individuals with lower scores.^5^ Independently, education has also been suggested as a proxy of cognitive reserve in both FTD^9-11, 47, 48^ and bvFTD specifically.^7^ Conversely, others have contested the association in bvFTD^49^ or restrict the this claim to certain disease phenotypes.^50^ Comparison of studies is difficult due to methodological heterogeneity. Furthermore, there are likely power issues in the published literature. In keeping with Harper et al 2022^24^ years of education was not associated with AAO in the present study. There is significant support for the brain reserve model in AD where gross anatomical measures such as head circumference and brain volume are identified proxies.^51-53^ To the best of our knowledge a proxy of brain reserve, as defined by Stern et al 2020,^39^ has yet to be established in FTD. Results from the present study alongside Harper et al 2022^24^ therefore provide evidence for the first proxy of brain reserve in FTD. The rapidity of decline in survival after disease onset in individuals possessing a right PCS is in accordance with reserve theories and the wealth of data in AD, whereby despite initial tolerance of disease burden, following phenoconversion individuals with higher reserve suffer a more rapid rate of clinical decline than individuals with low reserve.^25 40 54^ This phenomenon has been observed with respect to occupational attainment in bvFTD.^55^ Longitudinal analysis of clinical disease progression did not reach statistical significance in the present study due to powering, however a trend towards a more rapid rate of disease progression was observed in individuals with a right PCS, supporting the rapid decline theory following overload of compensatory mechanisms.

Right PCS presence was not associated with cortical thickness, a surrogate for cortical atrophy in this study. A disconnect between cortical atrophy and clinical disease expression is however described in bvFTD,^56 57^ suggesting cortical thickness may not provide an accurate representation of disease burden in all bvFTD sufferers.

The precise neurobiological substrate of reserve remains unknown although greater synaptic density, neuron quantity, brain size, advantageous metabolic properties and increased cerebral blood flow have all been suggested as potential structural and functional underpinnings.^5, 9, 47, 48^ Functionally, disruption of the salience network, an intrinsic resting state network anchored in the AC, is correlated with clinical severity in bvFTD.^49, 58, 59^ PCS presence has been shown to alter the functional architecture of the AC cortex at rest with hemispheres possessing a present PCS displaying enhanced connectivity.^60^ Furthermore, gyrification is considered to reflect the density of structural neural connectivity, with the degree of cortical folding partially pathway-specific dependent on mechanical tensions.^61-65^ As such, a salience network topographically and/or structurally altered by the presence of a right PCS may therefore possess resilience to bvFTD.

The right laterality of our findings is relevant for several reasons. The right dorsal ACC (dACC) is active unilaterally, early in decision making and monitoring of cognitive conflict.^66 67^ Thus, right but not left dACC could be more closely linked with development of core bvFTD symptoms. Secondly, the salience network is organisationally dominant in the right hemisphere^68, 69^ with multimodal structural and functional imaging studies^68, 70 69 68^ identifying stronger and broader intrinsic functional network couplings in the right compared to left dACC. Finally, VENs which are selectively targeted in bvFTD are more numerous in the right than left hemisphere.^71, 72^

A trend towards a later AAO in females with sporadic bvFTD^73^ and all cause bvFTD^74^ has been reported. Others have demonstrated better than expected executive function in females despite similar levels of atrophy to males.^74^ In keeping with previous work^24^ however we did not observe a direct association between sex and AAO in sporadic bvFTD. An interaction effect was observed with right PCS presence such that males with a present PCS had a later AAO than other subgroups. As reported by Illán-Gala et al 2021^74^ survival after AAO in the present study was similar in males and females.

This study is subject to limitations, importantly neuropathological diagnostic verification was available in only a minority of individuals. Furthermore, determination of AAO and assessment of clinical disease severity may be subject to bias. Finally, lifetime exposures with a potential impact on bvFTD onset including but are not limited to occupation, physical exercise, and dietary habits were not accounted for in this study but have been considered to impact upon cognitive reserve.^39, 75^

The effect of PCS on disease progression requires further study in a sufficiently powered cohort with neuropathological and longitudinal clinical and radiological data. The impact of PCS presence on disease expression and progression remains unstudied in a genetic bvFTD and is highly indicated given that therapeutic trials of disease-modifying therapies for bvFTD will likely be studied first in genetic cases. The present study identifies that gyrification in a region with a predilection to early and extensive pathological insult by a neurodegenerative disease provides resilience to clinical disease expression. Future study may explore the impact of relevant local gyrification patterns across the spectrum of the neurodegenerative diseases.

### Summary

Findings presented in the present study indicate an association between right PCS presence and disease expression and survival in sporadic bvFTD, providing evidence for the first proxy of brain reserve in FTD which may be important in the design of trials for future therapeutic approaches.

## Data Availability

All data produced in the present study are available upon reasonable request to the authors

## Acknowledgements

We express our gratitude toward all participating subjects, their next of kin, all personnel from the four studies contributing with material to this work and the funding bodies.

