## Supplemental File for "Anterior cingulate sulcation is associated with onset and survival in frontotemporal dementia"

**Materials and Methods**

**Participants**

**Penn FTDC**

Individuals with bvFTD from the Penn Frontotemporal Dementia Centre (Penn FTDC), The University of Pennsylvania, Pennsylvania, USA met clinical criteria for possible, probable, or definite bvFTD^1^ after multidisciplinary FTDC consensus conference discussion. Participants were included between 2003 and 2018. Exclusion criteria from Raskovsky et al 2011 were specifically applied as follows: Individuals with abrupt onset or step-wise progression of cognitive or behavioral symptoms; early anterograde memory difficulty; other primary neurodegenerative conditions (e.g. AD; Parkinson’s disease with early resting tremor); vascular disease (>Fazekas Stage 1 on MRI); hydrocephalus; head trauma; primary psychiatric conditions (e.g. major depression, schizophrenia, bipolar); medical conditions (e.g. metabolic, infectious), pacemaker or other cardiac device that would impact ability for MRI scanning were excluded. Participants who met criteria for bvFTD but had evidence of AD biomarkers based on reduced CSF t-tau/Aβ42/40 ratio ^2^ were excluded from all analyses. Ethical approval was given by the University of Pennsylvania Institutional Review Board (IRB).

**Amsterdam Dementia Cohort**

The Amsterdam Dementia Cohort ^3^ is a longitudinal study comprised of mixed retrospective and prospective data for individuals with Dementia of all types. All bvFTD patients were diagnosed by consensus by a multidisciplinary team after a standardized clinical work-up including a clinical consultation, neuropsychological testing, blood, CSF, brain imaging and EEG or MEG analysis. Genetic screening is offered to all FTD patients. If in diagnostic doubt, further clinical investigation and examination is offered depending on the differential diagnosis. In the case of FTD this includes a second evaluation by a neurologist together with a psychiatrist, FDG-PET, cognitive tests focusing on social cognition and/or follow-up, and more recently serum NfL. Participant inclusion began in 2000 and is currently ongoing. Specific inclusion- and exclusion criteria, diagnostic and neuropsychological procedure and imaging protocols have been described previously, ^3^. Additional exclusion criteria for the present study included: any significant neurological or psychiatric comorbidity, CSF biomarker analysis strongly suggestive of Alzheimer´s disease. The study was approved by the Medical Ethical Committee of the AmsterdamUMC, location VUmc, Amsterdam, Holland.

**MRI acquisition and Software**

**Penn FTDC**

Subjects were scanned on three different scanners: 3T Siemens Trio, 3T Prisma and I.5T Siemens Sonata. The following acquisition parameters were used: Trio (MPRAGE), three protocols: 3T Siemens Trio, protocol 1: axial plane, repetition time (TR) 1.62 ms, echo time (TE) 3.87 ms, inversion time (TI) 950 ms, flip angle (FA) 15°, voxel size 0.98×0.98×1 mm. 3T Siemens Trio: protocol 2: axial plane, TR x 1.62 ms, TE 3.09 ms, TI x 950 ms, FA 15°, voxel size 0.98×0.98×1 mm. 3T Siemens Trio, protocol 3: sagittal plane, TR 2.3 ms, TE 2.95 ms, TI 900 ms, FA 9°, voxel size 1.2×0.98×1 mm. 3T Prisma Fit (MPRAGE), two protocols, 3T Prisma, protocol 1: sagittal plane, TR 2.3 ms, TE 2.91 ms, TI 900 ms, FA 9°, voxel size 1.2x1.06x1.06mm. 3T Prisma, protocol 2: sagittal plane, TR 2.4 ms, TE 1.96 ms, TI 1020 ms, FA 8°, voxel size 0.8x0.8x0.8mm. 1.5T Seimens Sonata (MPRAGE): axial plane, TR 1900 ms, TE 4.38 ms, TI 1100 ms, FA 15°, voxel size 0.98×0.98×1 mm.

**Amsterdam Dementia Cohort**

Subjects were scanned on 8 different scanners: 1T Siemens Magnetom Impact, 1.5T Siemens Vision, 1.5T Siemens Sonata, 1.5T Siemens Avanto, 1.5T GE SignaHDxt, 3T GE SignaHDxt, 3T Toshiba Titan and a 3T Philips Ingenuity PET/MR system. The following acquisition parameters were used: 1T Siemens Magnetom Impact (MPRAGE): Coronal plane, repetition time (TR) 15 ms, echo time (TE) 7ms, inversion time (TI) 300 ms, flip angle (FA) 15°, voxel size 1×1×1.5 mm. 1.5T Siemens Vision (MPRAGE): coronal plane, TR 15 ms, TE 7ms, FA 8°, voxel size 0.98x0.98x1.5mm. 1.5T Siemens Sonata (MPRAGE): coronal plane, TR 2700 ms, TE 3.97 ms, TI 950 ms, FA 8°, voxel size 1×1×1.5 mm. 1.5T Siemens Avanto (MPRAGE): coronal plane, TR 2700 ms, TE 5.2 ms, TI 950 ms, FA 8°, voxel size 1×1×1.5 mm. 1.5T SignaHDxt (FSPGR): sagittal plane, TR 12.4 ms, TE 5.17 ms, TI 450 ms, FA 12°, voxel size 0.98×0.98×1.5 mm. 3T SignaHDxt (FSPGR): sagittal plane, TR 8 ms, TE 3 ms, TI 450, FA 12°, voxel size 0.98×0.98×1 mm. 3T Toshiba Titan (FFE): sagittal plane, TR 9.5 ms, TE 3.2 ms, TI 800 ms, FA 7°, voxel size 1×1×1 mm. 3T Philips Ingenuity PET/MR (TFE): sagittal plane, TR 8 ms, TE 4 ms, FA 12°, voxel size 1×1×1 mm.

**MRI pre-processing**

Cortical reconstruction and volumetric segmentation were performed on T1 3D MR images using FreeSurfer Software version 7.3.2 image analysis pipeline, (http://surfer.nmr.mgh.harvard.edu/). This procedure is described in prior publications including ^4^, listed at <https://surfer.nmr.mgh.harvard.edu/fswiki/FreeSurferMethodsCitation>. Briefly, whole-brain T1-weighted images underwent correction for intensity homogeneity, skull striping, and segmentation into grey and white matter with intensity gradient and connectivity among voxels. Cortical thickness was calculated as the distance between the grey matter and white matter boundaries (white matter surface) to grey matter and cerebrospinal fluid boundaries (pial surface) on the cortex of each hemisphere. Individuals’ cortices were anatomically parcellated and resampled to FresSurfer’s average surface map (fsaverage). A 10-mm full-width half-maximum Gaussian spatial smoothing kernel was applied to the surface maps. Reconstructed data sets were visually inspected for accuracy with quality control of each image performed by a single rater. Cortical thickness was successfully calculated in 176 individuals.

**Power Calculation**

Assuming a similar frequency of right PCS presence and relationship between right PCS presence and AAO as reported in our previous study^5^, where the mean AAO in individuals with a present right PCS was 63.77 and 60.38 in individuals with an absent right PCS, a power calculation was performed using a two-sample, one-sided t-test. A desired population of 173 individuals was identified.

Power calculation: Alpha 0.05. Power 0.8. Two-sample T-test with a one-sided hypothesis yields 74 individuals per group. Given the frequency of right PCS presence in the population was 0.57, 173 individuals with bvFTD shall be recruited in order to identify a minimum of 74 individuals per group.

**Paracingulate Sulcus Measurement and Classification Criteria**

Images were imported into MANGO (Multi-image Analysis GUI, v 4.0, <http://ric.uthscsa.edu/mango/mango.html>, The University of Texas Health Science Centre) software and prepared, aligning the x-axis in the sagittal plane with the bicommissural line (AC–PC). Further y- and z-axes rotational corrections were performed in order to ensure optimal orientation for analysis of PCS presence. Manual PCS classification was performed radiographically according to a protocol adapted from Garrison’s established protocol for PCS classification ^6^, used and described in Harper et al 2022^5^. An intra-rater agreement of 98.33%, *Cohens Kappa* 0.96 and an inter-rater agreement of 94.44%, *Cohens Kappa* 0.85 has been reported in reliability studies using this protocol ^7^. A binary sulcation classification was utilised where the PCS was categorized as either “present” (≥20 mm) or “absent” (≤19 mm) as is standard amongst PCS classification protocols.^6, 8-11^. The cingulate sulcus (CS) is identified 4 mm laterally from the midline (x = 0). The PCS is identified as the sulcus running predominantly horizontally, dorsal and parallel to the CS. The anterior limit of the PCS is identified as the point at which the sulcus begins to move posteriorly and parallel to the CS from an imaginary line perpendicular to the AC–PC line ^12^. The PCS is measured from this point using MANGO’s (Multi-image Analysis GUI, v 4.0, <http://ric.uthscsa.edu/mango/mango.html>, The University of TexasHealth Science Center) “Trace Line” function until its end point, the point where the sulcus is interrupted by a distinct predominantly vertical gyri deemed non-PC in nature. The PCS may fall outside of the first quadrant but must originate in the first quadrant on a sagittal plane where x = 0, y = 0 marks the point of the anterior commissure after images have been aligned in the AC-PC plane. The measurement protocol was modified in the present study and Harper et al 2022^5^, such that discontinuous PCS lacking an individual segment ≥20 mm in length were classified as “absent”. This decision was made as raters in this study found measurement of these structures to be inconsistent and unreliable in pre-study rater training. In addition, present PCS were required to be visible on four consecutive sagittal slices (a total depth of >4mm). Measurement was performed on the sagittal slice where the anteroposterior PCS length was shortest of the four longest consecutive sagittal slices where the PCS could be identified. This adaption meant that sulcation length was maintained throughout a depth of four sagittal slices which we believe to have increased the reliability of detection of a true PCS. Additionally, present PCS were subclassified as “prominent” where their length exceeded 40mm, as performed by others^6^ ^12^. Sulcation ratings were performed by two raters, LH and AS, who were blinded to individuals’ clinical and demographic data. Disagreement between raters was resolved by consensus.

**Results**

**Supplementary Fig. 1. Interaction Effect of Right Paracingulate presence and Sex on Age at Onset**

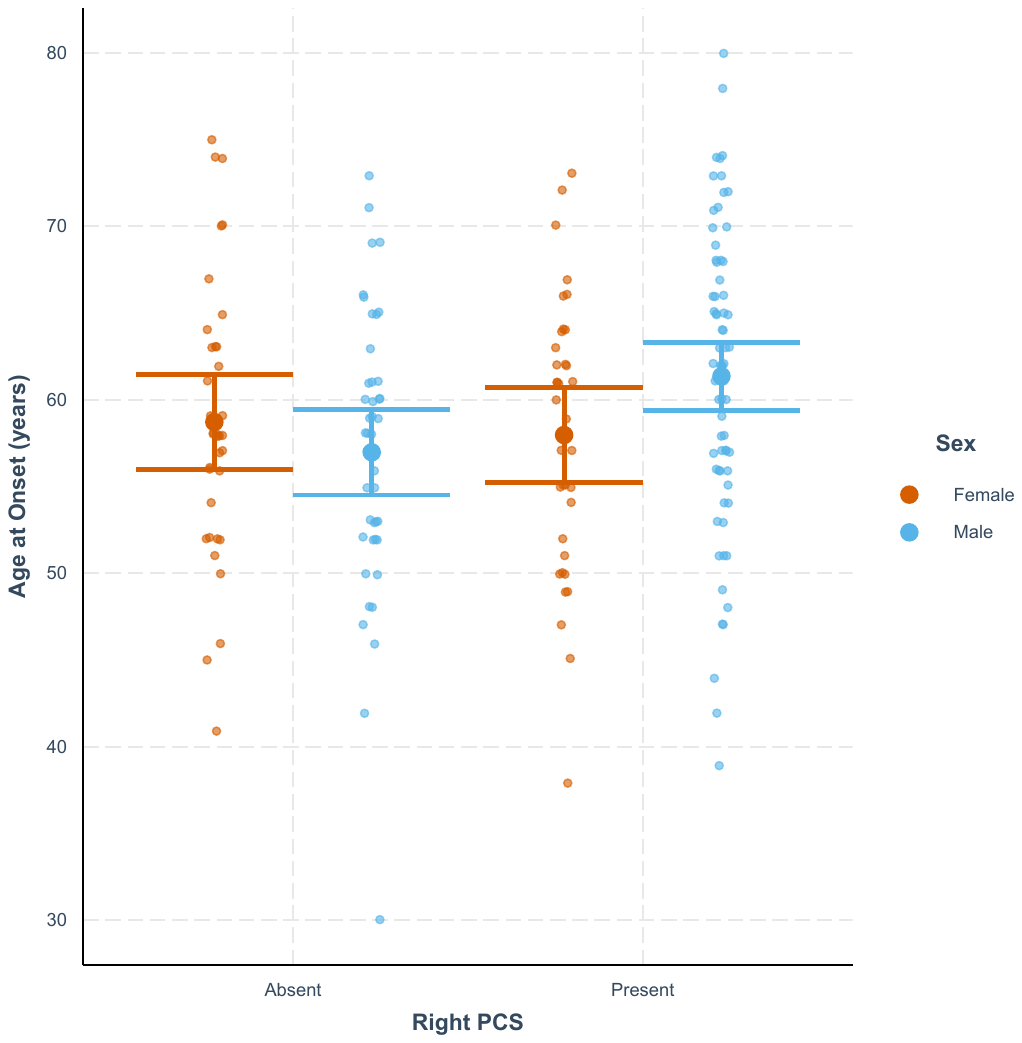

Linear regression with interaction effect between right PCS and Sex on AAO. Orange dots represent females. Blue dots represent males.

**Supplementary Fig. 2. Clinical Disease Progression in bvFTD by Right Paracingulate Sulcal Presence**

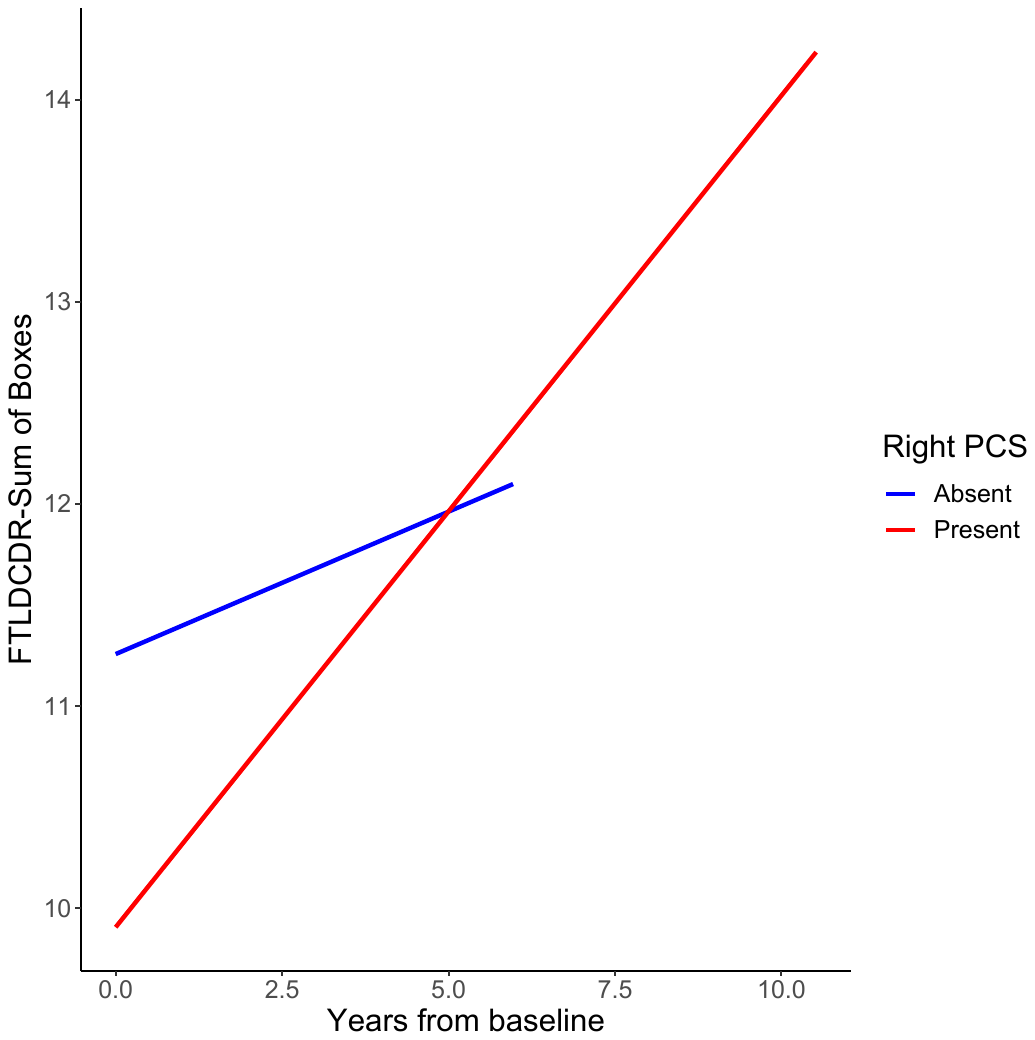

FTLD-CDR Sum of boxes scores (FTLDCDR-SB) were analyzed by fitting a linear mixed‐effects model with random intercepts and slopes with covariates education, age, handedness and sex. For illustrative purposes, we display the results of right PCS absent and present individuals by blue and red lines, respectively.

**Supplementary Table 1. Demographic and Results by Cohort and Right Paracingulate Sulcal Presence**

|  | **Entire Cohort** | | | | **Penn FTDC** | |  | **Amsterdam Dementia Cohort** | |  |  |
| --- | --- | --- | --- | --- | --- | --- | --- | --- | --- | --- | --- |
|  | Total | Right PCS absent | Right PCS present | p-value | Right PCS absent | Right PCS present | p-value | Right PCS absent | Right PCS present | *P*-value | Statistical Test |
| Participants, No. (%) | 186 | 80 (43) | 106 (57) |  | 36 (38) | 58 (62) |  | 44 (48) | 48 (52) |  |  |
| Age, mean (SD), years | 63.28 (8.32) | 62.29 (7.29) | 64.06 (9.02) | 0.18 | 61.6 (8.27) | 64.15 (9.12) | 0.13 | 62.72 (6.7) | 63.98 (9.03) | 0.23 | T-test (1-tail) |
| Age at Onset, mean (SD), years | 59.15 (8.4) | 57.76 (8.05) | 60.2 (8.54) | 0.024 | 56.63 (8.18) | 59.12 (8.29) | 0.08 | 58.68 (7.93) | 61.48 (8.74) | 0.06 | T-test (1-tail) |
| Age at Scan, mean (SD), years | 62.27 (8.59) | 61.63 (7.61) | 62.76 (8.54) | 0.36 | 60.61 (8.46) | 61.72 (9.17) | 0.28 | 62.45 (6.84) | 64.02 (9.30) | 0.18 | T-test (1-tail) |
| Sex | 113M: F73 | 44M: 36F | 69M: 37F | 0.21 | 23M: 13F | 36M: 22F | 1 | 21M: 23F | 33M: 15F | 0.07 | Chi-Squared |
| [Education^a^, means (SD), years](javascript:;) | 13.34 (3.79) | 12.96 (4.21) | 13.62 (3.44) | 0.26 | 16.56 (2.62) | 15.47 (2.74) | 0.06 | 9.8 (2.42) | 11.34 (2.79) | 0.01 | T-test (2-tailed) |
| [Handedness](javascript:;), No |  |  |  | 0.82 |  |  | 0.81 |  |  | 0.25 | Chi-Squared |
| Right | 146 | 62 | 84 |  | 30 | 51 |  | 32 | 33 |  |  |
| Left | 12 | 4 | 8 |  | 4 | 5 |  | 0 | 2 |  |  |
| Ambidextrous | 5 | 2 | 3 |  | 2 | 2 |  | 0 | 1 |  |  |
| Unknown | 23 | 12 | 11 |  | 0 | 0 |  | 12 | 12 |  |  |
| MMSE, mean (SD) | 24.18 (5.35) | 24.49 (4.58) | 23.97 (5.84) | 0.52 | 24.46 (4.79) | 23.12 (6.73) | 0.27 | 24.51 (4.44) | 25.04 (4.32) | 0.31 | T-test (2-tailed) |
| FTLD CDR Sum of boxes, mean (SD) | 7.51 (3.83) | 7.98 (4.12) | 7.13 (3.56) | 0.21 | 9.87 (4.35) | 8.43 (3.23) | 0.23 | 6.22 (3.76) | 6.32 (3.55) | 0.33 | T-test (2-tailed) |
| FTLD CDR Global scores, mean (SD) | 1.59 (0.72) | 1.67 (0.73) | 1.53 (0.7) | 0.25 | 1.89 (0.57) | 1.71 (0.6) | 0.3 | 1.57 (0.78) | 1.41 (0.74) | 0.59 | T-test (2-tailed) |
| Deceased^b^, No. (%) | 112 (0.6) | 43 (0.54) | 69 (0.65) |  | 15 (0.42) | 36 (0.62) |  | 28 (0.64) | 33 (0.69) |  |  |
| Age at Death, mean (SD), years | 67.19 (8.86) | 67.62 (7.38) | 66.94 (9.66) | 0.7 | 69.14 (8.87) | 66.75 (9.28) | 0.41 | 66.56 (6.16) | 67.23 (10.43) | 0.79 | T-test (2-tailed) |

Demographic and Results data for entire population and according to cohort, Penn FTDC (The University of Pennsylvania Frontotemporal Degeneration Centre, Pennsylvania, USA) and Alzheimer center of the VU University Medical Center, Amsterdam (Amsterdam Dementia Cohort).^3^ Standard deviation (SD). Sex data: Male (M). Female (F). ^a^ Education, years data available for 94/94 Penn FTDC and 88/92 Amsterdam Dementia Cohort. ^b^ *Survival data was available for 185/186 individuals.* Mini Mental State Examination (MMSE). Frontotemporal Lobar Degeneration-modified Clinical Dementia Rating (FTLD-CDR)*^13^*, Sum of boxes (Sum) and Global score (Global). Hemispheric Paracingulate Sulcus (Hemispheric PCS), present = PCS length ≥20mm, prominent = PCS length ≥40mm. Frequency. T-tests and Chi-Squared tests were performed to evaluate differences in continuous and nominal data, respectively*.*

**Supplementary Table 2. Right Hemisphere Regional Cortical Thickness Differences**

| **Right Hemisphere** | |  |  |  |  |
| --- | --- | --- | --- | --- | --- |
| **Region** | **Centrode Coordinate** | **Centrode -log10 (p-value)** | **p-value** | **Anatomical Location** | **Cortical Thickness in Right PCS Present Individuals vs Right PCS absent Individuals** |
| 1 | 10.71, 13.11, 43.30 | -1.9 | 0.01 | Right Mid Cingulum | Reduced |
| 2 | 9.06, 40.67, 25.23 | -1.4 | 0.04 | Right Anterior Cingulum | Reduced |
| 3 | 30.92, 26.75, 7.23 | -1.6 | 0.03 | Right Insula | Reduced |
| 4 | 32.40, 10.88, 29.67 | -1.4 | 0.04 | Right Frontal Inferior Operculum | Reduced |
| 5 | 27.34, 11.14, 45.24 | -1.5 | 0.03 | Right Frontal Mid | Reduced |
| 6 | 37.82, -28.85, 56.45 | 1.3 | 0.05 | Right Postcentral | Increased |
| 7 | 29.83, -48.48, 45.66 | -1.5 | 0.03 | Right Parietal Inf | Reduced |
| 8 | 9.28, -27.26, 70.74 | -1.5 | 0.03 | Right Paracentral Lobule | Reduced |
| 9 | 11.59, -34.92, 76.12 | -1.6 | 0.03 | Right Postcentral | Reduced |
| 10 | 17.63, 38.34, -18.19 | -1.5 | 0.03 | Right Frontal Superior Orbital | Reduced |

Right Hemisphere regions with differences in cortical thickness prior to cluster correction for multiple analyses. Threshold *P* <0.05.

**Supplementary Table 3. Left Hemisphere Regional Cortical Thickness Differences**

| **Left Hemisphere** | |  |  |  |  |
| --- | --- | --- | --- | --- | --- |
| **Region** | **Centrode Coordinate** | **Centrode -log10 (p-value)** | **p-value** | **Anatomical Location** | **Cortical Thickness in Right PCS Present Individuals vs Right PCS absent Individuals** |
| 1 | -11.39, 24.84, 25.40 | -2 | 0.01 | Left Anterior Cingulum | Reduced |
| 2 | -8.14, -47.94, 18.34 | -1.5 | 0.03 | Left Precuneus | Reduced |
| 3 | -8.46, -53.81, 47.05 | 1.5 | 0.03 | Left Precuneus | Increased |
| 4 | -29.00, -51.51, 49.17 | -1.4 | 0.04 | Left Inferior Parietal | Reduced |
| 5 | -55.58, -40.77, 44.05 | 2.3 | 0.005 | Left Inferior Parietal | Increased |
| 6 | -56.10, -56.50, -2.44 | 1.4 | 0.04 | Left Mid Temporal | Increased |
| 7 | -62.98, -30.12, -16.38 | 1.6 | 0.03 | Left Inferior Temporal | Increased |
| 8 | -57.07, -17.93, 21.20 | 1.6 | 0.03 | Left Postcentral | Increased |
| 9 | -58.05, 5.06, 26.36 | 1.3 | 0.05 | Left Precentral | Increased |
| 10 | -51.96, -28.16, -29.32 | 1.5 | 0.03 | Left Inferior Temporal | Increased |
| 11 | -53.57, -22.23, -26.21 | 1.5 | 0.03 | Left Inferior Temporal | Increased |
| 12 | -41.07, -1.75, -38.51 | 1.5 | 0.03 | Left Inferior Temporal | Increased |
| 13 | -56.69, -2.68, -28.50 | 1.3 | 0.05 | Left Inferior Temporal | Increased |

Left Hemisphere regions with differences in cortical thickness prior to cluster correction for multiple analyses. Threshold *P* <0.05.
